# Exome re-analysis driven by deep phenotyping increases diagnostic yield

**DOI:** 10.1101/2022.09.13.22279858

**Authors:** Atanu Kumar Dutta, Niladri Sekhar Bhunia, Rohit Bhowmik, Nihar Ranjan Mishra, Rimjhim Sonowal, Kalyan Goswami, Anbu Kayalvizhi C

**Author notes:** Corresponding author: Atanu Kumar Dutta, Department of Biochemistry, Second Floor, Medical College Block, All India Institute of Medical Sciences, Kalyani, NH - 34 Connector Basantapur, Saguna, Kalyani, 741245, West Bengal, India. **Email address of all other authors:**; Niladri Sekhar Bhunia -; Rohit Bhowmik -; Nihar Ranjan Mishra -; Rimjhim Sonowal -; Kalyan Goswami –; Anbu Kayalvizhi C. **Ethical statement:** The Institute Ethics Committee had given ethics waiver for the current work on 31th August 2022. **Consent statement:** Written informed consent was obtained from the participants/ their legal representatives regarding their participation in this research study as well as to have the results published for the wider scientific community without revealing their identity. **Contribution:** AKD, KG – conceiving the study; NSB, RB, NRM, RS – Paediatric evaluation of cases; AKD, NSB – Clinical Genetics evaluation of cases including facial dysmorphology and database searching; AKD, AKC – Exome re-analysis including CNV analysis; All authors – manuscript preparation and finalization.

## Abstract

**Objective:** To explore if clinician driven systematic reanalysis of “negative” exome sequencing data of patients with a strong suspicion of a Mendelian disease by a Clinical Geneticist using deep phenotyping can increase the diagnostic yield.

**Methods:** Ten patients were deep phenotyped using Gestalt matching (Face2Gene) and OMIM database searching (Phenomizer) to build a list of candidate genes of interest. The Variant Call Format (vcf) files for individual patients were obtained from the sequencing laboratory and reanalyzed in the wANNOVAR server. The annotated list of variants was queried for the genes of interest. Copy number variant analysis was performed using ExomeDepth (v1.1.10) R package.

**Results:** A homozygous pathogenic mutation in the *NPHP3* gene (c.2805C>T) was identified in one child with Renal-Hepatic-Pancreatic Dysplasia and a multiexon deletion in the *CUL7* gene was identified in another child with 3M syndrome.

**Conclusions:** Systematic re-analysis of exome sequencing data by a Clinical Geneticist can increase the diagnostic yield with minimal additional cost.

## Introduction

With a rapid decline in sequencing cost many Paediatricians are now routinely prescribing Exome Sequencing for children with suspected Mendelian disorders. However, even after this, the diagnosis remains elusive in about 40% of the cases (1). Literature suggests the range of additional diagnostic yield of 6% - 22% can be obtained through reanalysis of the Exome data (1). Exome re-analysis also led to a higher possibility of offering a prenatal diagnosis. The factors leading to increased diagnostic yield on re-analysis include i) additional literature evidence; ii) additional phenotypic features develop or are detected, iii) trio/ additional family member Exome sequencing, iv) improved sequencing, v) improved bioinformatic analysis pipelines, vi) identification of new genes responsible for Mendelian diseases, vii) research collaboration leading to the delineation of functional effect, viii) formation of international clinical collaborative efforts such as PhenoDB, GeneMatcher, and VariantMatcher (2), ix) ability to perform copy number variation analysis in Exome data (1). While noncoding or structural variants can be important causes in the remaining cases the lack of detailed genotype-phenotype correlation can be likely cause of missing the diagnosis. A number of recent studies had highlighted the clinical utility of Exome rea-analysis in diverse phenotypes which integrated workflows involving Machine Learning and Deep Phenotyping. Periodic, cost-effective reanalysis may benefit patients and their families and physicians (3). However, a recent systematic review and meta-analysis have revealed that though next-generation sequencing data reanalysis can improve diagnostic yield, the optimal time to reanalysis and the impact of AI-based tools could not be determined with confidence (3). Here we intended to explore the utility of this approach in the Indian context with the resources available in a typical teaching hospital.

## Methods

Ten children/families with strongly suspected Mendelian disease but a “negative” Exome report who were referred to the Paediatric Genetic Clinic were prospectively included in this study from August 2021 to August 2022. They were evaluated by a Paediatrician and a Clinical Geneticist. Facial dysmorphology analysis was performed using the DeepGestalt algorithm of the Face2Gene website (3). In addition, the OMIM database was searched for the identified HPO terms by the Phenomizer API (4). A list of candidate genes was extracted from the Phenomizer output. The clinical details of the children are presented in table 1. The Variant Call Format (vcf) files for individual patients were obtained from the sequencing laboratory and reanalyzed in the wANNOVAR server (5). The annotated variant output was analyzed in the wAANOVAR web interface for candidates’ genes of interest. Copy number variant analysis was performed using ExomeDepth (v.1.1.10) R package (6). The identified variants were visualized using the Integrated Genomic Viewer in the proband and parents (where available) to rule out sequencing artefacts (7). Ethics committee of All India Institute of Medical Sciences, Kalyani waived ethical approval for this work. Written informed consent was obtained from the participants/ their legal representatives regarding their participation in this research study as well as to have the results published for the wider scientific community without revealing their identity.

**Table 1.** Characteristics of the study subjects

| Family number | Proband initial clinical diagnosis at referral | Proband Initial Exome result | Exome re-analysis result | Reason for diagnosis |
| --- | --- | --- | --- | --- |
| 1 | Polycystic Kidney Disease | *BICC1*: p.Lys744Gln | **Renal-Hepatic-Pancreatic Dysplasia**<br>**NPHP3: c.2805C>T (p.Gly935(=))** | Update in medical literature |
| 2 | Aicardi-Goutières syndrome | Nil | Unchanged | - |
| 3 | Vici Syndrome | Oculocutaneous albinism type II (*OCA2*: c.1970G>T in heterozygous state) | Unchanged | - |
| 4 | Wilson Disease | Nil | Unchanged | - |
| 5 | KBG Syndrome (sibling also affected) | Nil | Unchanged | - |
| 6 | Loeys-Dietz Syndrome (sibling also affected) | Nil | Unchanged | - |
| 7 | Achondroplasia (sibling also affected) | Nil | **3 M syndrome (CUL7 gene multiexon deletion)** | Patient has phenotype update |
| 8 | Gitelman Syndrome | Nil | Unchanged | - |
| 9 | KAKUT | *KMT2D*: c.7219C>A | Unchanged | - |
| 10 | Kabuki Syndrome | Nil | Unchanged | - |

## Results

The clinical details of the included families are listed in table 1. For families seven and ten causal pathogenic variants were identified. For the rest of the families no causal variant could be identified (table 1).

### Family 1

#### Renal-Hepatic-Pancreatic Dysplasia

A child born of a non-consanguineous marriage presented with antenatally detected bilateral cystic kidneys, hepatic cyst, scalp haemangioma, hypertension, Ostium Secundum atrial septal defect, and hyperpigmented patch. As the child was clinically suspected to suffer from polycystic kidney disease the child was prescribed Exome sequencing to confirm the diagnosis. However, no pathogenic or likely pathogenic variant was reported. Subsequently, other family members also underwent Exome sequencing without revealing the causal etiology in the child. We were consulted by the couple for planning for the next pregnancy. We searched the Phenomizer using the following query terms: Polycystic kidney dysplasia (HP:0000113); Laryngomalacia (HP:0001601); Haemangioma (HP:0001028); Hepatic cysts (HP:0001407) and Atria septal defect (HP:0001631). The significant genetic associations (p-value 0.002 to 0.008) were the following: *PRKCSH/ LRP5/ SEC63; PKD1; NEK8/ NPHP3; PKHD1; ANKS6; DYNC2H1/ NEK1* and *KAT6B*. We retrieved the raw data of the trio exome and analyzed it in the wAANOVAR server to look for variants specifically in these genes. Of the 24,919 variants detected in the sequence data of the proband the only variant with extremely low population allele frequency (0.00000813 in gnomAD exomes) in any of the candidate genes was *NPHP3*: c.2805C>T (p.Gly935(=)) [GrCh38]. This variant was present in a homozygous state in the proband and a heterozygous state in the parents. This was consistent with an autosomal recessive mode of inheritance. This variant was overlooked in the prior automated analysis being a synonymous variant. The same variant was recently reported in six unrelated families from Oman, the US, the UK, and Saudi Arabia in children with Renal-hepatic-pancreatic dysplasia 1 (OMIM #208540) (8). This variant was missed in the initial Exome analysis in these families as well. The authors also performed an analysis of the whole blood derived RNA and identified both wild-type and aberrantly spliced mRNA confirming a predicted activation of a novel splice donor inside exon 20 of the *NPHP3* gene leading to fusion of exon 20, intron 20, and exon 21. The altered splicing led to frameshift and truncation of the final protein product. However, the author could not replicate the study in the RNA from the liver and kidney due to nonavailability. This variant leads to the activation of a cryptic exonic splice donor leading to frameshift and shorter *NPHP3* transcript. This highlights the need to keep the rare synonymous variants during the filtering steps. Recent studies have shown that these variants can lead to altered mRNA splicing, slowed initiation and speed of translation, altered mRNA structure, and stability, co-translational protein misfolding disrupted microRNA binding sites, and altered post-translational modifications (9). Cryptic splice sites have been estimated to account for ∼10% of cases with rare genetic disorders (10). The models for splicing disruptions include - i) additional splice donor site, ii) additional splice acceptor site, iii) skipping of upstream exon/s, iv) skipping of downstream exon/s, and v) retention of the intron. Blood RNA analysis can help to resolve VUS and increase the diagnostic rate (11). (Further clinical details of the index family can be obtained from the corresponding author.)

### Family 7

#### 3M syndrome

Two siblings born of a non-consanguineous marriage presented to the genetic clinic with proportionate short stature and dysmorphism. The elder sibling had Exome sequencing done previously with a clinical suspicion of Achondroplasia but no pathogenic causative variant was identified. Their height and weight were less than the third centile but the head circumference of the younger sibling was in the 25^th^ centile. They had a triangular face, midface retrusion, thick eyebrows, depressed nasal bridge, full lips, pointed chin, square shoulders, short neck, flat feet, prominent heels, and generalized joint hypermobility. There was no history of developmental delay or intellectual disability. They had slender long bones with diaphyseal constriction and flared metaphyses. The vertebra was tall with irregular end plates. Metacarpal bones have high cortical thickness. A facial dysmorphology assessment was carried out using Face2Gene. Based on the following diagnostic handles: Proportionate Short stature HP:3508; Depressed nasal bridge HP:0005280; Diaphyseal thickening HP:0005019; Decreased body weight HP:0004325; Increased vertebral height HP:0004570; Anteverted nares HP:0000463; Pointed chin HP:0000307; Autosomal recessive inheritance HP:0000007; Triangular face HP:0000325 and Prominent forehead HP:0011220 the 3M syndrome was the most probable diagnosis (p-value 0.0014). The genes of interest were *CCDC8, OBSL1* and *CUL7*. We then retrieved the raw data of the exome and analyzed it in the wAANOVAR server to look for variants specifically in these genes. However, there was no pathogenic or likely pathogenic variant identified. Because of strong clinical suspicion of 3M syndrome, we performed copy number variant analysis of the Exome data using the ExomeDepth (v1.1.10) R package. Which revealed homozygous deletion of 10 exons (4811 bases) in the *CUL7* gene (chr6: 43045587 – 43050398) [GRCh38]. (Further clinical details of the index family can be obtained from the corresponding author.)

## Conclusions

Exome re-analysis driven by deep phenotyping and systematic database searching allowed us to diagnose two out of ten patients. Our approach was broadly similar to Wang *et al* (9). Pedigree with more than one system involved had a higher rate of diagnosis from Exome. Exome reanalysis can be requested by the i) clinician, ii) clinical laboratory, or iii) the patient (12).

Renal-Hepatic-Pancreatic Dysplasia (RHPD), a genetically diverse ciliopathy was first described by Ivemark *et al* in 1959. The liver, Kidney, and pancreas undergo progressive cystic dysplasia and fibrosis leading to early death. To date homozygous or compound heterozygous pathogenic variants in the *NPHP3* and *NEK8* genes have been attributed to RHPD1 and 2 respectively. Nephrocystin (*NPHP3*) is part of a ciliary protein complex required for organogenesis. RHPD1 is allelic to Nephronophthisis 3 (milder variant) and Meckel syndrome 7 (severe variant). Nephronophthisis is the commonest inherited cause of ESRD in children. Hypomorphic NPHP3 mutation leads to cystic kidney disease phenotype whereas complete loss leads to situs inversus, congenital heart disease, and embryonic lethality. NPHP3 is responsible for both early embryonic development as well as maintenance of morphologic integrity throughout life (13). Cysts developed due to disrupted planar orientation of cells due to malrotation of mitotic spindle complex leading to nondirected cell division (14). Nephronophthisis is the most common monogenic cause of End Stage Renal Disease in children accounting for up to 15% of this population (14). However, to date, only 40% of patients receive a genetic diagnosis (15).

CUL7 appears to be the major gene responsible for 3M syndrome accounting for 77.5% of cases while OBSL1 mutations account for 16.3% (16). CUL7 accounts for 70% of cases of 3M syndrome. The distinctive features of 3M syndrome related to the CUL7 gene include full eyebrows, hypospadias, small testis, and joint hypermobility. The siblings in our report had full eyebrows and joint hypermobility (17). This is the first report of 3M syndrome due to exonic deletion of the *CUL7* gene. This case highlights the requirement of routine CNV analysis of the Exome data especially when the clinical suspicion is high.

Common mistakes leading to missed diagnosis in Exome analysis include i) poor sequence quality, ii) presence of alternate contigs in the reference genome, iii) calling variants only in the exome capture targets, iv) improper reference pools for CNV calling, v) not updating gene definitions as per RefSeq, vi) not visually inspecting the data in IGV, vii) missing variants other than non-synonymous variants, viii) missing hidden compound heterozygous variant, ix) missing mosaicism, x) missing structural chromosomal variants, xi) missing highly prevalent disease-causing variants, xii) misleading phenotypic information, xiii) non-Mendelian inheritance, xiv) presence of pseudogenes and xv) gene copies (18). In nondiagnostic Exomes apart from re-analysis the following approaches may help to resolve the diagnosis – i) short read genome sequencing, ii) long-read genome sequencing, iii) RNA sequencing, iv) case matching, v) using variant aware Pan-genome, vi) phenotype driven metabolomics, vii) epigenomics, viii) proteomics, ix) functional studies in model systems, x) integration of multi-omics data (19).

Automated analysis of the Electronic Health Record by extracting the HPO terms (4) and then automated periodic Exome re-analysis (20) has also become more and more standardized. To conclude systematic re-analysis of Exome data in our clinical genetics set up lead to 20% higher diagnostic yield with minimal additional cost.

#### What this Study Adds

Periodic re-analysis of Exome supplemented by deep phenotyping increase the diagnostic yield

## Data Availability

All data produced in the present work are contained in the manuscript

## Acknowledgements

To the participating families.

To the support staff of the Department of Paediatrics, AIIMS Kalyani & MedGenome Laboratories.

